# Clustering Lifestyle Risk Behaviors among Vietnamese Adolescents and Roles of School: A Bayesian Multilevel Analysis of Global School-Based Student Health Survey 2019

**DOI:** 10.1101/2021.03.26.21254389

**Authors:** Khuong Quynh Long, Hoang Thi Ngoc-Anh, Nguyen Hong Phuong, Tran Thi Tuyet-Hanh, Kidong Park, Momoe Takeuchi, Nguyen Tuan Lam, Pham Thi Quynh Nga, Le Phuong-Anh, Le Van Tuan, Tran Quoc Bao, Ong Phuc Thinh, Nguyen Van Huy, Vu Thi Hoang Lan, Hoang Van Minh

## Abstract

**Background:** Adolescence is a vulnerable period for many lifestyle risk behaviors. In this study, we aimed to 1) examine a clustering pattern of lifestyle risk behaviors; 2) investigate roles of the school health promotion programs on this pattern among adolescents in Vietnam.

**Methods:** We analyzed data of 7,541 adolescents aged 13–17 years from the 2019 nationally representative Global School-based Student Health Survey, conducted in 20 provinces and cities in Vietnam. We applied the latent class analysis to identify groups of clustering and used Bayesian 2-level logistic regressions to evaluate the correlation of school health promotion programs on these clusters. We reassessed the school effect size by incorporating different informative priors to the Bayesian models.

**Findings:** The most frequent lifestyle risk behavior among Vietnamese adolescents was physical inactivity, followed by unhealthy diet, and sedentary behavior. Most of students had a cluster of at least two risk factors and nearly a half with at least three risk factors. Latent class analysis detected 23% males and 18% females being at higher risk of lifestyle behaviors. Consistent through different priors, high quality of health promotion programs associated with lower the odds of lifestyle risk behaviors (highest quality schools vs. lowest quality schools; males: Odds ratio (OR) = 0·67, 95% Highest Density Interval (HDI): 0·46 – 0·93; females: OR = 0·69, 95% HDI: 0·47 – 0·98).

**Interpretation:** Our findings demonstrated the clustering of specific lifestyle risk behaviors among Vietnamese in-school adolescents. School-based interventions separated for males and females might reduce multiple health risk behaviors in adolescence.

**Funding:** The 2019 Global School-based Student Health Survey was conducted with financial support from the World Health Organization. The authors received no funding for the data analysis, data interpretation, manuscript writing, authorship, and/or publication of this article.

## Introduction

Non-communicable diseases (NCDs) are responsible for almost 70% of all deaths worldwide [1] and becoming more common among youth nowadays [2]. Health risk behaviors are activities that increase the risk of diseases or injuries if doing with enough frequency or intensity [3]. These behaviors begin in early life that affects health both at that time and later years [4]. According to WHO, two-thirds of premature deaths in adults are associated with childhood risk behaviors, such as 81% of youth aged 11–17 years were physical inactivity and 11·7% were heavy drinking [5]. Most NCDs share predisposing risk factors, for example, people suffered from diabetes and cancer as concurrent exposure to unhealthy diets, physical inactivity, and harmful use of tobacco and/or alcohol [5]. These factors are unlikely to isolate, but, instead, typically cluster and interact to exponentially elevate the risks of NCDs [6]. Findings in more than 300,000 adolescents from 89 countries showed that 82·4% of them exposed to several NCDs risk factors, including unhealthy diet and physical inactivity; or unhealthy diet and cigarette smoking [7].

Adolescence is a critical period for developing and forming a healthy lifestyle [8], and also a vulnerable period for several unhealthy behaviors which might continue into maturity [9]. Adolescents tend to be involved in more than one problem due to shared linkages of such behaviors [10-13]. School going adolescents spent at least six hours a day at school on their social, physical, and intellectual development. The school setting becomes an ideal place for targeted lifestyle programs that equipped healthy behaviors for adolescents before their transition into adulthood [14]. Some previous studies suggested that school health programs can reduce the prevalence of health risk behaviors among youth [15]. Although school environments can affect student health [16], important evidence gaps about the roles of schools in alleviating health risk factors in adolescents remain. Understanding this gap is essential to improve adolescent health and reduce subsequent disease burden in adulthood.

In Vietnam, a surged prevalence of NCDs in recent years is associated with the country’s remarkable economic growth and lifestyle changes [17]. In 2016, NCDs contributed to 73% of total deaths due to dietary risks, tobacco smoke, alcohol use, and physical inactivity [18]. The Vietnam national NCDs strategy 2015-2025 focuses on preventing NCDs among adolescents aimed to reduce smoking prevalence below 3·6%; overweight and obesity prevalence below 10% in this population [19].

Existing literature about the clustering of lifestyle risk behaviors in low- and middle-income countries (LMICs) is currently limited. However, most of the prior studies used descriptive techniques to analyze risk behaviors individually using arbitrary cut-off points [20] or determining high-risk groups via the ratio of observed-to-expected prevalence [21, 22]. Also, limited evidence has focused on the roles of school health promotion programs. In Vietnam, to the best of our knowledge, no study published on clustering of risk behaviors to date. In this paper, we aimed to: 1) describe a clustering pattern of six lifestyle risk behaviors (smoking, drinking, physical inactivity, sedentary behaviors, low fruit/vegetable intake, and unhealthy diet) using latent class analysis (LCA) - an analytical technique to find underlying patterns of such behaviors; 2) to investigate the roles of the school health promotion programs on these behavior patterns using Bayesian multilevel models with different informative priors.

## Methods

### Study population and survey design

The 2019 Global School-based Student Health Survey (GSHS) is a population-based survey of school-going adolescents aged 13-17 years, which has been conducted over 101 countries [23], providing data on different aspects of adolescent behaviors and protective factors to help countries develop suitable adolescent health programs and policies [23]. In this study, we used the nationally representative sample of Vietnam, which was conducted from May to December 2019 across 20 provinces and cities in Vietnam.

### Questionnaire development

The 2019 Vietnam GSHS employed a set of global self-administered questions that adapted to the local socio-cultural context. A panel including four language and content experts was established to validate the content. First, the experts translated forward the original English questionnaire into Vietnamese, then backward translation from Vietnamese into English. The original English version was compared to backward translation to check consistency. Face validity was assessed from a pilot study through pre-testing the translated questionnaire in 120 students in one secondary school and one high school in Hanoi city, Vietnam. The validated questionnaire has two main components: socio-demographics (age, gender, living status, and place of residence) and health behaviors (alcohol use, tobacco use, dietary behavior, self-perceived body mass index (BMI) status, illicit drug use, mental health, physical activity, sexual behaviors, and violence and unintentional injuries).

### Sample size and sampling procedures

We used a two-stage cluster sampling method to recruit adolescents. At the first stage, we chose schools based on probability proportional to size method, then following by the selection of classes using the simple random sampling technique at the second stage. Two classes for each secondary school (grades 8 and 9) and three classes for each high school (grades 10, 11, and 12) were selected. All students who were 13–17 years old and had Vietnamese citizenship in selected classes were eligible to participate. The sample included 7,796 students in 210 classes from 81 schools. The school response rate of 96·4% and the student response rate of 97·0% made up the overall response rate of 93·5%.

Before data collection, school administrators assisted to distribute written assent forms to the parents/guardians in selected classes. Parents/guardians were asked to return the forms regardless of their agreement. During data collection days, we only recruited students with parental permission to participate into the study. Trained researchers briefed and guided on the self-administered questionnaire, then students marked their responses on a separate computer scan-able answer sheet. The time to complete the questionnaire was maximum 30 minutes. All completed anonymous sheets were sealed in envelopes to ensure confidentiality.

### Variables

#### Lifestyle risk behaviors

##### Smoking

We defined the current smoking status by asking students the number of days that they used any tobacco products in the past 30 days. The variable response was dichotomized into *“0 days”* and *“at least 1 day”*.

##### Alcohol consumption

We assessed the current drinking by a question on the number of days that students drank at least one standard drink of alcoholic beverage (∼14gram of pure alcohol) in the past 30 days. The variable response was categorized as *“0 days”* and *“at least 1 day”*.

##### Physical inactivity

Physical activity is defined as any body movement generated by the contraction of skeletal muscles that raises energy expenditure above resting metabolic rate [24].

We asked students a question on the number of days that they had at least 60 minutes of moderate-to-vigorous-intensity physical activity in the past seven days. We defined the physical inactivity if respondents did not meet the WHO recommendation for physical activity [25]. Thus students were categorized as “physical inactivity” if they did not report doing at least an average of 60 minutes per day of moderate-to-vigorous intensity physical activities in seven days a week.

##### Sedentary behavior

Sedentary behaviors are defined as any waking behaviors that consumed an energy expenditure equal to or below 1·5 metabolic equivalents. Screen time and sitting time are usually the two main indicators used to quantify the time devoted to sedentary behaviors [26, 27].

To assess the sedentariness, we asked students a question on the hour spent on sitting and watching television, playing computer games, using social media, or doing other sitting activities, on an average day in the past seven days. We classified respondents as having sedentary behavior if they spent on more than two hours a day doing those activities [28].

##### Low fruit/vegetable intake

We asked students questions on the average daily frequency intake of fruits and vegetables in the past 30 days. We defined the low fruit/vegetable intake if they did not consume both fruits and vegetables at least two times per day [29].

##### Unhealthy diet

We defined unhealthy diet students if they drank carbonated soft drinks at least one time per day during the 30 days before the survey or ate fast-food at least one day during the 7 days before the survey [29].

#### School health promotion programs

We asked students whether they received training for five soft-skills (Yes/no), including (1) the benefits of eating more fruits and vegetables, (2) signs of depression and suicidal behaviors, (3) problems associated with drinking alcohol, (4) problems associated with using drugs, and (5) benefits of physical activity. We then aggregated these variables at the school level and constructed the proxy measurement of school health promotion programs quality by using PCA to compute a composite index. This composite index was categorized into tertiles [30], with higher tertiles indicating better school program quality.

#### Other covariates

The other covariates include demographic characteristics (age, place of residence, living with mother/father), body mass index–Z score, parental monitoring, peer-relationship (number of close friends), mental health (loneliness and worrying), and truancy. The details on variable definitions are provided in **Supplemental materials S2**.

### Data analysis

After removing missing values on covariates (3·3%), which was assumed to be missing at random, the complete case sample analyzed in this paper was 7,541 (3,495 males and 4,046 females).

#### Sampling weights calculation

We calculated the sampling weights reflecting the likelihood of sampling each student and to reduce bias by compensating for different patterns of nonresponse. The sampling weights were given by

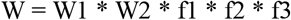

In which, W1 is the inverse of the probability of selecting the school; W2 is the inverse of the probability of selecting the classroom within the school; f1 is a school level nonresponse adjustment factor; f2 is a student-level nonresponse adjustment factor; f3 is a post-stratification adjustment factor calculated by rural/urban and grade.

#### Descriptive analysis

We used frequencies and percentages to summarize categorical variables, mean and standard deviation (SD) to describe continuous variables. We adjusted all estimations for the complex survey design, including sampling weights, clustering, and stratification. To describe the combinations of six lifestyle risk behaviors, we used the UpSet diagrams [31], which visualize complex intersections of a lifestyle risk behaviors matrix where the rows represent different sets of combinations and the columns represent the number of students who had these combinations.

#### Lifestyle risk behaviors clustering and related factors

We developed an analysis framework (**Figure 1**) that includes two steps to analyze the clustering and related factors.

**Figure 1:** Analysis framework. *Note: LCA: Latent Class Analysis; PCA: Principal Component Analysis*.

In the first step, we used LCA to identify homogeneous unobservable subgroups of lifestyle risk behaviors. LCA is a statistical method for identifying unmeasured class membership among subjects using categorical or continuous observed variables [32]. To explore the potential number of latent classes, we tested the LCA with a different number of classes (i.e., from two to five classes). We determined the final number of classes based on the interpretability of the class memberships [32]. The details in LCA results were provided in the **Supplemental materials S3**. In the second step, we fitted a series of Bayesian 2-level random intercept logistic regressions with students at level-1, and school at level-2 to evaluate the effects of factors in school and student levels on latent class memberships obtained from LCA in the first step. Multilevel modeling was used to account for the nature of hierarchical data i.e., students nested in schools.

We applied the vague priors (i.e., flat prior for model parameters). Model 1 was a null model with no independent variable, Model 2 included the school-level factors (X_1j_), and Model 3 controlled for both school-level and student-level covariates (X_2ij_).

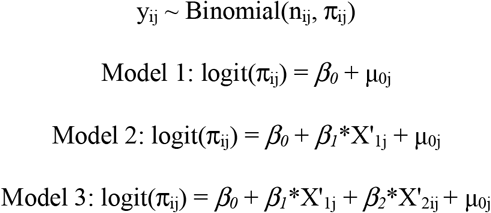

The priors for model parameters were specified as:

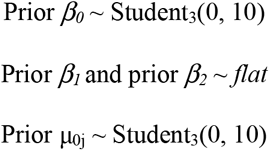

Where μ_0j_ is the school-specific residual that presents the deviate of each school from the median log odds of high risk of lifestyle behaviors. For each model, we calculated the variance partition coefficient (VPC) and the proportion of variance explained by the added factors (i.e., % explained). Since logistic regression has a variance of 3·29 [33], the VPC is defined as VPC = σ^2^_μ0_/(σ^2^_μ0_ + 3·29). To obtain % explained, we subtracted the variance of the simpler model to the model with more terms and then converted to percentages. The school-level and student-level covariates included in the model were determined by the conceptual framework of effects of school health promotion programs on lifestyle risk behaviors that was based on the conceptual framework developed by Akseer et al [34]. The conceptual framework is shown in **Supplemental materials S4**.

We presented the median of posterior distributions as odds ratio (OR) with 95% highest density interval (95% HDI). We also calculated the probability of the posterior distribution of school effect size (i.e., ORs) less than the cut-offs of 1, 0·9, and 0·8 as well as Bayes factors (BF). The BF is the ratio of the likelihood of a specific hypothesis to the opposite hypothesis [35]. Therefore, in this case, the larger value of the BF indicates the stronger evidence of the hypothesis that the school effect size was lower than the cut-offs.

#### Sensitivity analysis

We conducted sensitivity analyses to test whether the school effects remain consistent across different priors. We constructed two priors to reflect two degrees of belief that the school health promotion programs had either no impact (*equivocal prior*) or positive impact (*optimistic prior*) on lifestyle risk behaviors of students. We assumed that prior distributions of school effects were the normal distribution (i.e., *β*_*1*_ ∼ Normal(μ, σ^2^)).

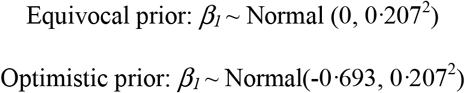

Details of sensitivity analyses were provided in **Supplemental materials S4**.

We fitted the Bayesian multilevel models using Hamiltonian Monte Carlo (HMC) algorithm. With the expected effect sample sizes were greater than 10000, we generated four HMC chains in parallel, the iterations of 8000, the burn-in period of 1000, and the thinning of 2. The convergence of HMC chains was diagnosed by trace plot and the Gelman-Rubin coefficient [36]. We used Stata v16 SE (StataCorp, College Station, TX, USA) to clean the data and conduct descriptive analysis; R (version 4·0·0, R Foundation for Statistical Computing, Vienna, Austria) to conduct the LCA using package *poLCA* [37] and the Bayesian multilevel models using package *brms* that interfaces with Stan [36].

### Ethical approval

All procedures performed in this study followed the ethical standards of the Institution Review Board of Hanoi University of Public Health (IRB decision No. 421/2019/YTCC-HD3, dated: 06/08/2019).

## Results

### Participant characteristics

**Table 1** describes the characteristics of the study participants. Our weighted sample represented a population of 5,400,584 adolescents. More than half of students were females (54·1%) and nearly two thirds of them lived in rural areas. Most of them lived with their mother or father and had more than two close friends. The proportion of students felt lonely and anxious in the past 12 months was 11·7% and 4·8% in males and 12·8% and 7·2% in females, respectively. The percentages of students truanted in the past 30 days were 17·1% and 13·0% among males and females.

**Table 1:** Participants’ characteristics.

| Characteristics | Males |  | Females |  |
| --- | --- | --- | --- | --- |
|  | n<sup>a</sup> | %<sup>b</sup> | n<sup>a</sup> | %<sup>b</sup> |
| **N weighted = 5,400,584** | 3495 | 45.9 | 4046 | 54.1 |
| **Student-level variables** |  |  |  |  |
| **Age** |  |  |  |  |
| 13 | 426 | 15.5 | 569 | 17.3 |
| 14 | 667 | 28.6 | 830 | 32.2 |
| 15 | 795 | 21.7 | 828 | 18.1 |
| 16 | 744 | 16.3 | 884 | 15.9 |
| 17 | 863 | 18.0 | 935 | 16.5 |
| **BMI-Z score, mean (SD)** |  | -0.4 (1.3) |  | -0.5 (1.0) |
| **Place of residence** |  |  |  |  |
| Rural | 1717 | 62.7 | 2041 | 64.2 |
| Urban | 1778 | 37.3 | 2005 | 35.8 |
| **Living with mother/father** |  |  |  |  |
| No | 583 | 17.2 | 669 | 17.5 |
| Yes | 2909 | 82.8 | 3376 | 82.5 |
| **Parental monitoring, mean (SD)** |  | 5.7 (2.3) |  | 5.4 (2.2) |
| **Number of close friends** |  |  |  |  |
| 0 | 347 | 8.4 | 417 | 9.0 |
| 1 | 386 | 10.3 | 665 | 15.2 |
| 2 | 447 | 12.9 | 699 | 17.6 |
| ≥3 | 2269 | 68.3 | 2218 | 58.2 |
| **Loneliness** |  |  |  |  |
| No | 3026 | 88.3 | 3438 | 87.2 |
| Yes | 468 | 11.7 | 608 | 12.8 |
| **Anxiety** |  |  |  |  |
| No | 3295 | 95.2 | 3700 | 92.8 |
| Yes | 194 | 4.8 | 344 | 7.2 |
| **Truancy** |  |  |  |  |
| No | 2782 | 82.9 | 3418 | 87.0 |
| Yes | 614 | 17.1 | 544 | 13.0 |
| **School health promotion programs** |  |  |  |  |
| **Percentage of students taught about, mean (SD)** |  |  |  |  |
| Benefits of eating more fruits and vegetables |  | 82.3 (10.1) |  | 82.9 (9.6) |
| Signs of depression and suicidal behaviors |  | 20.8 (8.4) |  | 21.4 (8.1) |
| Problems associated with drinking alcohol |  | 70.9 (9.7) |  | 71.5 (9.2) |
| Problems associated with using drugs |  | 84.4 (6.8) |  | 85.2 (6.7) |
| Benefits of physical activity |  | 81.2 (11.7) |  | 81.8 (11.1) |
| **Percentage of students taught > 3 soft skills, mean (SD)** |  | 45.4 (11.1) |  | 45.9 (10.3) |
| **School quality proxy** |  |  |  |  |
| 1<sup>st</sup> tertile | 1232 | 25.4 | 1295 | 21.4 |
| 2<sup>nd</sup> tertile | 1204 | 34.4 | 1385 | 35.7 |
| 3<sup>rd</sup> tertile | 1059 | 40.2 | 1366 | 42.9 |

<sup>a</sup>Unweighted frequency, <sup>b</sup>Weighted percentage

More than 70% of students were taught about the benefits of eating more fruits and vegetables, problems associated with drinking alcohol, drug consequences, and benefits of physical activity, but only about 20% of them received training about signs of depression and suicidal behaviors. Less than half of students achieved three such modules together (45·4% in males and 45·9% in females). Results from PCA analysis showed that more than 40% of students studied in high-quality schools.

### Distribution of lifestyle risk behaviors

**Figure 2.** shows the profile of lifestyle risk behaviors among in-school adolescents in Vietnam. The physical inactivity was the most frequent lifestyle risk behavior, followed by unhealthy diet and sedentary behavior. The percentage of smoking and drinking were higher among males than females (4·4% vs. 1·0%, and 24·7% vs. 20·0%, respectively). In contrast, females were more inactive than males, with a high prevalence of physical inactivity (90·1% vs. 77·9%) and sedentary behavior (47·8% vs. 37·3%).

**Figure 2:** Distribution of lifestyle risk behaviors among in-school adolescents in Vietnam. *Note: A: Percentage of lifestyle risk behaviors among males and females; B: Distribution of lifestyle risk behavior co-occurrence among males and females*.

Almost all students had at least one risk factor (96·8% in males and 98·5% in females). Many students had a combination of two (34·2% in males and 34·6% in females) and three factors (27·3% in males and 31·5% in females). Only <1% students had none of the six risk factors. Males and females equally distributed in each category of the number of risk factors.

**Figure 3.** describes the frequent combinations formed by six lifestyle risk behaviors. The most frequent cluster in both sexes was physical inactivity and unhealthy diet, with the proportion of 15·7% and 16·7% in males and females, respectively. The second most frequency was physical inactivity, sedentary behavior, and unhealthy diet in females, while physical inactivity in males. The combination of four risk factors physical inactivity, sedentary behavior, low fruit/vegetable intake, and unhealthy diet was the third most common combination.

**Figure 3:** UpSet diagrams for combinations of lifestyle risk behaviors. *Note: The combinations that have proportion less than 1% are not shown*.

### Cluster of lifestyle risk behaviors

Based on the criteria of interpretability from LCA analysis, we decided to choose two latent classes as the outcome for the clustering of lifestyle risk behaviors modeling. We named the two classes based on the distribution of each risk behavior in each class (i.e., higher and lower risk class of lifestyle behaviors). **Table 2** presents the distribution of lifestyle risk behaviors among participants by two class memberships. We detected 23·1% of males and 18·0% of females were at higher risk cluster of lifestyle behaviors. Details on LCA results were in the **Supplemental material S3**.

**Table 2:** Distribution of lifestyle risk behaviors among in-school adolescents in Vietnam, by two class memberships.

| Lifestyle health risk behaviors | Males |  | Females |  |
| --- | --- | --- | --- | --- |
|  | Lower risk cluster<br>(n = 2579, 76.9%) | Higher risk cluster<br>(n = 916, 23.1%) | Lower risk cluster<br>(n = 3250, 82.0%) | Higher risk cluster<br>(n = 796, 18.0%) |
| Smoking, % | 0.0 | 19.1 | 0.0 | 5.7 |
| Alcohol consumption, % | 4.1 | 93.5 | 2.8 | 98.3 |
| Physical inactivity, % | 78.1 | 77.2 | 90.1 | 90.1 |
| Sedentary behavior, % | 31.7 | 55.8 | 43.5 | 67.1 |
| Low fruit/vegetable intake, % | 30.6 | 34.9 | 29.1 | 35.3 |
| Unhealthy diet, % | 61.8 | 81.8 | 63.9 | 83.1 |
| **Number of health risk behaviors, %** |  |  |  |  |
| 0 | 4.2 | 0.0 | 1.8 | 0.0 |
| 1 | 23.1 | 0.1 | 16.6 | 0.0 |
| 2 | 41.6 | 9.5 | 41.2 | 4.8 |
| 3 | 24.6 | 36.4 | 31.2 | 32.9 |
| 4 | 6.5 | 37.8 | 9.3 | 41.8 |
| 5 | 0.0 | 14.6 | 0.0 | 19.1 |
| 6 | 0.0 | 1.6 | 0.0 | 1.4 |

### Role of the school health promotion programs on high-risk cluster of lifestyle behaviors

**Table 3** shows factors related to a high level of lifestyle risk behaviors in Vietnamese male and female adolescents. After controlling for student-level covariates, students in high-quality schools (3rd tertile) were less likely to have high level of lifestyle risk behaviors than those in low-quality schools (1st tertile) (Males: OR = 0·67, 95% HDI: 0·46 – 0·93; Females: OR = 0·69, 95% HDI: 0·47 – 0·98).

**Table 3:** Bayesian multivariable models of factors related to high-risk cluster of lifestyle behaviors among in-school adolescents in Vietnam.

|  | Model 1 |  | Model 2 |  | Model 3 |  |
| --- | --- | --- | --- | --- | --- | --- |
|  | Males<br>OR<br>(95% HDI) | Females<br>OR<br>(95% HDI) | Males<br>OR<br>(95% HDI) | Females<br>OR<br>(95% HDI) | Males<br>OR<br>(95% HDI) | Females<br>OR<br>(95% HDI) |
| **Fixed part** |  |  |  |  |  |  |
| **Intercept** | 0.30 (0.25–0.35) | 0.20 (0.17–0.24) | 0.35 (0.26–0.47) | 0.23 (0.17–0.30) | 0.24 (0.14–0.43) | 0.13 (0.08–0.23) |
| **School quality proxy (Ref: 1<sup>st</sup> tertile)** |  |  |  |  |  |  |
| 2nd tertile |  |  | 1.08 (0.73–1.62) | 1.18 (0.77–1.78) | 0.98 (0.70–1.38) | 1.10 (0.77–1.59) |
| 3rd tertile |  |  | **0.58 (0.39–0.88)** | **0.64 (0.42–0.94)** | **0.67 (0.46–0.93)** | **0.69 (0.47–0.98)** |
| **Student-level covariates** |  |  |  |  |  |  |
| **Age (Ref: 13)** |  |  |  |  |  |  |
| 14 |  |  |  |  | 1.08 (0.75–1.57) | **1.54 (1.08–2.22)** |
| 15 |  |  |  |  | 1.34 (0.93–1.95) | 1.48 (0.99–2.22) |
| 16 |  |  |  |  | **2.10 (1.41–3.06)** | **2.36 (1.58–3.55)** |
| 17 |  |  |  |  | **3.40 (2.32–5.00)** | **2.24 (1.51–3.40)** |
| **BMI-Z score** |  |  |  |  | 1.03 (0.96–1.10) | 0.94 (0.86–1.03) |
| **Place of residence (Ref: Rural)** |  |  |  |  |  |  |
| Urban |  |  |  |  | 1.18 (0.90–1.58) | 1.30 (0.96–1.76) |
| **Living with mother/father (Ref: No)** |  |  |  |  |  |  |
| Yes |  |  |  |  | **0.78 (0.63–0.98)** | 0.85 (0.68–1.05) |
| **Parental monitoring** |  |  |  |  | **0.93 (0.90–0.97)** | **0.93 (0.89–0.97)** |
| **Number of close friends (Ref: 0)** |  |  |  |  |  |  |
| 1 |  |  |  |  | 1.15 (0.78–1.61) | 1.02 (0.74–1.42) |
| 2 |  |  |  |  | 1.00 (0.69–1.40) | 1.22 (0.90–1.71) |
| ≥3 |  |  |  |  | 1.15 (0.86–1.53) | 1.17 (0.89–1.57) |
| **Loneliness (Ref: No)** |  |  |  |  |  |  |
| Yes |  |  |  |  | **1.34 (1.04–1.70)** | **1.53 (1.21–1.92)** |
| **Worrying (Ref: No)** |  |  |  |  |  |  |
| Yes |  |  |  |  | 1.20 (0.84–1.72) | **1.68 (1.27–2.21)** |
| **Truancy (Ref: No)** |  |  |  |  |  |  |
| Yes |  |  |  |  | **2.23 (1.82–2.74)** | **2.01 (1.50–2.49)** |
| **Random part** |  |  |  |  |  |  |
| Variance estimate | 0.46 (0.29–0.72) | 0.50 (0.29–0.72) | 0.40 (0.25–0.64) | 0.43 (0.27–0.71) | 0.23 (0.12–0.40) | 0.28 (0.15–0.48) |
| VPC (%)<sup>§</sup> | 12.4 | 13.3 | 11.0 | 11.8 | 6.6 | 7.9 |
| % Explained<sup>†</sup> | -- | -- | 12.5 | 12.2 | 42.9 | 36.0 |

HDI: Highest density interval; OR: Odds ratio
Model 1: A null 2-level random effects model, with students at level-1, and school at level-2; Model 2: Model 1 + main predictors (i.e., School quality proxy); Model 3: Model 2 + all student-level covariates; <sup>§</sup>VPC calculated as: $[\sigma^2_{\mu 0}/(\sigma^2_{\mu 0} + 3.29)]*100$ ; <sup>†</sup>% Explained calculated as: $[(\sigma^2_{\text{Model N}} - \sigma^2_{\text{Model N} + 1})/\sigma^2_{\text{Model N}}]*100$

In the base model without independent variable (Model 1), most variation of lifestyle risk behaviors clustering attributed to the student level; the school-level accounted for 12·4% and 13·3% of the variation among males and females, respectively. The inclusion of school-level factor explained ∼12% the between-school variation of lifestyle risk behaviors clustering, while student-level covariates explained further 42·9% and 36·0% of the variation.

### Sensitivity analysis

**Figure 4** and **Supplemental materials S4** provide the sensitivity analysis of the effects of school health promotion programs on students’ behaviors. These effects were different due to different priors, however, yielded the same conclusion: high-quality schools were associated with lower odds of high level of lifestyle risk behaviors in both males and females. Among males, the proportion of posterior distribution less than the cutoff of OR = 1 was from 96·8% (BF = 30·3) in equivocal prior to 100% (BF > 10000) in the optimistic prior. Among females, all results favored the hypothesis that the proportion of posterior distribution was less than the cutoff of OR = 1, with a minimum of 94·8% (BF = 18·1) in the equivocal prior. However, in equivocal prior, only 56·4% the posterior distribution of the effects of school quality on students’ behaviors was below the cutoff of OR = 0.8 among males; this figure for females was 49·7%.

**Figure 4:** Prior and posterior distributions the effects of school health promotion programs on high-risk cluster of lifestyle behaviors among in-school adolescents in Vietnam. *Note: The red dash lines represent the three priors: Vague prior: ∼ N(0, 1002); Equivocal prior:* *∼ N (0, 0·2072); Optimistic prior: ∼ N (−0·693, 0·2072). In each scenario, the posterior distributions represent the the effect of school promotion programs quality on lifestyle risk behavior clustering (i*.*e*., *the ORs of school promotion programs quality tertile 3 and tertile 2 vs. tertile 1 – reference group). The probability of the posterior distributions less than the cut-offs of 1, 0·9, and 0·8 (represented by three vertical lines) are shaded in a lighter gray color*. *All models were adjusted for age, body mass index Z-score, place of residence, living with mother/father, parental monitoring, number of close friend, loneliness, worrying, and truancy*.

### Other factors related to high-risk cluster of lifestyle behaviors

In both sexes, parental monitoring is associated with lower odds of lifestyle risk behaviors. In contrast, older students, those who felt lonely, or did truancy were more likely to have a higher level of lifestyle risk behaviors than those who did not. We also found that worrying was associated with higher lifestyle risk behaviors among females (**Table 3**).

## Discussion

This is the first study to investigate the prevalence, clustering pattern of six major lifestyle risk behaviors of NCDs (smoking, drinking, physical inactivity, sedentary behavior, low fruit/vegetable intake, and unhealthy diet) and its determinants among school-going adolescents aged 13-17 years in the nationwide scope in Vietnam. We found that lifestyle risk behaviors were common in Vietnamese adolescents. Among all NCDs’ risk factors, physical inactivity constituted the highest proportion in both males and females, followed by unhealthy diet and sedentary behavior intake. Most of students had a cluster of at least two risk factors and nearly a half with at least three risk factors. These factors tend to be clustered with the common patterns among physical inactivity and unhealthy diet in both sexes and sedentary behavior in females. The school health promotion programs quality is associated with lower the odds of lifestyle risk behaviors. Our study provides important empirical evidence to guide health promotion, education, and interventional programs of these lifestyle risk behaviors.

Previous studies have demonstrated the health risk behaviors among adolescents, however, we cautioned our comparisons due to variations in investigating different risk factors, definitions and cut-off points; and differing in targeted populations. Compared to a study in 2538 Malaysian school-going adolescents that employed the same definitions for smoking, alcohol use, sedentary behavior, and low fruit/vegetable intake, Vietnamese ones had a higher prevalence of alcohol use (24·7% in Vietnamese males and 20·0% in Vietnamese females vs. 6·6% in Malaysian males and 3·5% in Malaysian females) but lower levels of smoking, sedentary behavior, and low fruit/vegetable intake [22]. However, compared to another study conducted among 3990 Brazilian adolescents, Vietnamese adolescents had a similar prevalence of drinking alcohol (∼25%) but lower in smoking (4·4% in males and 1% in females in the present study vs 5·7%) [21].

Clustering multiple lifestyle risk behaviors are prevalent in Vietnamese adolescents. The prevalence of simultaneous occurrences of at least two risk behaviors was lower than that reported among Malaysian adolescents (∼60% vs. 83%) [22], Canadian children and adolescents (64%) [38] and Brazilian adolescents (53·8% in males and 71·1% in females) [21]. In concordance with previous literature, an physical inactivity, sedentary behavior, and unhealthy diet were prone to cluster either alone or together with other risk behaviors in both sexes [39, 40]. Physical inactivity, sedentary behavior, and unhealthy diet are three common obesogenic characteristics. The mechanism of clustering these obesogenic behaviors among adolescents is complex [41]. Prolonged physical inactivity and sedentary behavior are associated with an unhealthy diet by increasing consuming junk foods while doing sedentary activities such as sitting, watching television, or using a computer. This disrupts energy balance by increasing time for recreational physical activity that in turn can promote fat accumulation [42]. Physical inactivity and sedentary behavior combined with an unhealthy diet is negatively associated with health and wellbeing among adolescents and has been recognized as important lifestyle risk behaviors [43]. The clustering of health risk behaviors follow Jessor’s problem-behavior theory that adolescents engaged in one problem behavior tend to be involved in other problems due to the shared linkages of such behaviors in the social ecology [10]. We recommend that joint interventions on multiple risk behaviors can be more effective than individual approaches to tackle these synergistic clustering effects.

We observed more profound clustering effects of lifestyle risk behaviors in Vietnamese females than in males, and such findings corroborated current evidence [21, 22, 38]. Although generally known as lower risk-takers [44], adolescent females who engaged in one high-risk behavior tend to be more involved in others than their male counterparts. Furthermore, they have higher levels of perceived stress [45], a tendency to ruminate, and feelings of helplessness than males [46] that could predispose them to engage in dysfunctional coping measures [47]. Previous evidence indicated that adolescent females were more likely to be influenced by psychosocial motives [45, 46] and therefore more likely to engage in unhealthy risk behaviors [47]. As such, in-depth investigations into the actual causal factors could probably shed some light on the significant determinants of the clustering of risk behaviors among Vietnamese females. This finding implicated the separated design of public health interventions for males and females are needed. Our evidence supports the idea that the school health promotion programs are important for promoting and supporting healthy lifestyles among adolescents [48-50]. Previous studies indicated that school-based interventions are effective and feasible to improve healthy dietary habits, physical activity levels, and weight control [51]. Schools also reach a wide range of children over a considerable amount of time. Therefore, enhancing the school health promotion programs could be a prominent way to improve children’s health and well-being [52, 53]. In Vietnam, education on preventing lifestyle risk behaviors is not prerequisites in school; they are considered as elective modules; therefore, the quality, as well as content, might be not sufficient. It is noticed that despite of the high prevalence of studying an individual module for preventing lifestyle risk behavior, only less than half of students completed three or more such modules. This lack of joint knowledge on preventing lifestyle risk behaviors might lead to the pervasiveness of lifestyle risk behaviors in our sample. We highlight a need for the required courses that not only training on the harms of each risk behavior separately but also emphasize the connection of these behaviors [54].

Regarding sensitivity analysis, we used the Bayesian approach to analyze the effect of school health promotion programs quality on students’ behaviors among different scenarios of belief. All the scenarios, even in the equivocal view which is conservative to our belief, yield the same conclusion, it is more certain and confident to claim the robustness of findings. Another advantage of using Bayesian approach is its ability to measure the effect size. In our study, we found the strong evidence that high tertile of school health promotion programs quality had positively affect students’ behaviors (i.e., ORs < 1) compared to low tertile of quality, however, this effect became uncertain when compared to the cutoff of 0·8, (i.e., only 56·4% and 49·7% the posterior distribution was below the cutoff of OR = 0·8 among males and females in the equivocal prior, respectively). Together, our findings suggest that the effect size of school health promotion programs quality on students’ behaviors probably ranges from 0·8 to 1.

Strengths of our study included using robust statistical technique to investigate clusters of NCDs risk factors and its determinants among adolescents using large nationally representative sample. We used latent class analysis that was able to identify the underlying pattern of lifestyle risk behaviors co-occurrence and Bayesian regression models with different informative priors to investigate the relationship between these cluster and school health promotion programs. This is the first study to investigate the current status of lifestyle risk behaviors in Vietnam. Most current evidence investigating clusters of risk factors was done on adult populations or in high-income countries; not many studies conducted in lower-middle income countries. Despite its innovative approach, the present analysis has certain limitations. First, information on lifestyle risk behaviors was self-reported, which may lead to under or overestimation. Second, due to the availability of the data, the cutoff point for low fruit/vegetable intake cannot follow WHO recommendation. Third, we are unable to collect various aspects of both the school and student’s levels, such as parental education, income, social-economic status, which maybe potential predictors of clustering risk behaviors. Fourth, due to the cross-sectional nature of the data, we could not confirm the causal inferences. Fifth, although most adolescents in Vietnam are in school [55], our study focused only in-school adolescents, which may not represent for those who are not in school.

## Conclusion

Our findings demonstrated the clustering of specific combinations of lifestyle risk behaviors among Vietnamese in-school adolescents. School-based interventions separated for males and females might reduce multiple health risk behaviors in adolescence. A deeper understanding of clustered patterns may lead to developing new and comprehensive interventions to prevent the burden of NCDs in Vietnamese adolescents.

## Data Availability

The datasets analyzed for the current study are not publicly available but are available upon reasonable request.

## Acknowledgments

The authors would like to thank all students who participated in the study as well as the individuals and institutions that made this study possible: Departments of Education and Training from selected provinces, principals, and teachers from 81 schools who helped us to prepare for the data collection. Associate Prof. Tran Dac Phu and Dr. Truong Dinh Bac from the General Department of Preventive Medicine – Ministry of Health; Mr. Nguyen Thanh De and Mr. Le Manh Hung from the Ministry of Education and Training; Ms. Leanne Riley from World Health Organization; Ms. Veronica Lea, Ms. Curtis Blanton and Mr. Timothy McManus from the US CDC; and Mr. Cao Huu Quang from Hanoi University of Public Health.

## References

1. World Health Organization. Noncommunicable diseases: WHO; https://www.who.int/health-topics/noncommunicable-diseases#tab=tab_1. Accessed 25 Nov 2020.

2. Baker R, Taylor E, Essafi S, Jarvis JD, Odok C. Engaging young people in the prevention of noncommunicable diseases. Bull World Health Organ. 2016;94(7):484.

3. Steptoe A. Health Behavior and Stress*. In: Fink G, editor. Encyclopedia of Stress (Second Edition). New York: Academic Press; 2007. p. 262–6.

4. El Achhab Y, El Ammari A, El Kazdouh H, Najdi A, Berraho M, Tachfouti N, et al. Health risk behaviours amongst school adolescents: protocol for a mixed methods study. BMC public health. 2016;16(1):1209.

5. World Health Organization. WHO Global Coordination Mechanism on the Prevention and Control of NCDs: NCD and Youth: WHO; https://www.who.int/global-coordination-mechanism/ncd-themes/ncd-and-youth/en. Accessed 25 Nov 2020.

6. Arena R, Guazzi M, Lianov L, Whitsel L, Berra K, Lavie CJ, et al. Healthy lifestyle interventions to combat noncommunicable disease-a novel nonhierarchical connectivity model for key stakeholders: a policy statement from the American Heart Association, European Society of Cardiology, European Association for Cardiovascular Prevention and Rehabilitation, and American College of Preventive Medicine. European heart journal. 2015;36(31):2097–109.

7. Uddin R, Lee EY, Khan SR, Tremblay MS, Khan A. Clustering of lifestyle risk factors for non-communicable diseases in 304,779 adolescents from 89 countries: A global perspective. Preventive medicine. 2020;131:105955.

8. Lee RL, Loke AY, Wu CS, Ho AP. The lifestyle behaviours and psychosocial well-being of primary school students in Hong Kong. Journal of clinical nursing. 2010;19(9-10):1462–72.

9. Ness AR, Maynard M, Frankel S, Smith GD, Frobisher C, Leary SD, et al. Diet in childhood and adult cardiovascular and all cause mortality: the Boyd Orr cohort. Heart. 2005;91(7):894–8.

10. Jessor R. Risk behavior in adolescence: A psychosocial framework for understanding and action. Journal of Adolescent Health. 1991;12(8):597–605.

11. Nascente FM, Jardim TV, Peixoto MD, Carneiro CS, Mendonça KL, Póvoa TI, et al. Sedentary lifestyle and its associated factors among adolescents from public and private schools of a Brazilian state capital. BMC public health. 2016;16(1):1177.

12. Oyeyemi AL, Ishaku CM, Oyekola J, Wakawa HD, Lawan A, Yakubu S, et al. Patterns and Associated Factors of Physical Activity among Adolescents in Nigeria. PLoS One. 2016;11(2):e0150142–e.

13. Urrutia-Pereira M, Oliano VJ, Aranda CS, Mallol J, Solé D. Prevalence and factors associated with smoking among adolescents. Jornal de pediatria. 2017;93(3):230–7.

14. DeBar LL, Ritenbaugh C, Aickin M, Orwoll E, Elliot D, Dickerson J, et al. Youth: a health plan-based lifestyle intervention increases bone mineral density in adolescent girls. Archives of pediatrics & adolescent medicine. 2006;160(12):1269–76.

15. Bonell C, Wells H, Harden A, Jamal F, Fletcher A, Thomas J, et al. The effects on student health of interventions modifying the school environment: systematic review. Journal of Epidemiology and Community Health. 2013;67(8):677.

16. Bonell C, Parry W, Wells H, Jamal F, Fletcher A, Harden A, et al. The effects of the school environment on student health: a systematic review of multi-level studies. Health & place. 2013;21:180–91.

17. Nguyen TT, Hoang MV. Non-communicable diseases, food and nutrition in Vietnam from 1975 to 2015: the burden and national response. Asia Pacific journal of clinical nutrition. 2018;27(1):19–28.

18. World Health Organization. United Nations Interagency Task Force on the prevention and control of noncommunicable diseases. Joint mission, Vietnam. Geneva: WHO Press; 2016.

19. Ministry of Health of Vietnam. National strategy for the prevention and control of communicable diseases, period 2015-2025. Hanoi, Vietnam 2015.

20. Bener A, Ghuloum S, Abou-Saleh MT. Prevalence, symptom patterns and comorbidity of anxiety and depressive disorders in primary care in Qatar. Social psychiatry and psychiatric epidemiology. 2012;47(3):439–46.

21. Dumith SC, Muniz LC, Tassitano RM, Hallal PC, Menezes AM. Clustering of risk factors for chronic diseases among adolescents from Southern Brazil. Preventive medicine. 2012;54(6):393–6.

22. Teh CH, Teh MW, Lim KH, Kee CC, Sumarni MG, Heng PP, et al. Clustering of lifestyle risk behaviours and its determinants among school-going adolescents in a middle-income country: a cross-sectional study. BMC public health. 2019;19(1):1177.

23. World Health Organization. Global school-based student health survey (GSHS): WHO; https://www.who.int/ncds/surveillance/gshs/en. Accessed 25 Nov 2020.

24. Caspersen CJ, Powell KE, Christenson GM. Physical activity, exercise, and physical fitness: definitions and distinctions for health-related research. Public health reports (Washington, DC : 1974). 1985;100(2):126–31.

25. World Health Organization. Physical activity 2020. https://www.who.int/news-room/fact-sheets/detail/physical-activity. Accessed 25 Nov 2020.

26. Gibbs BB, Hergenroeder AL, Katzmarzyk PT, Lee IM, Jakicic JM. Definition, measurement, and health risks associated with sedentary behavior. Medicine and science in sports and exercise. 2015;47(6):1295–300.

27. Thivel D, Tremblay A, Genin PM, Panahi S, Rivière D, Duclos M. Physical Activity, Inactivity, and Sedentary Behaviors: Definitions and Implications in Occupational Health. Front Public Health. 2018;6:288-.

28. Chaput J-P, Willumsen J, Bull F, Chou R, Ekelund U, Firth J, et al. 2020 WHO guidelines on physical activity and sedentary behaviour for children and adolescents aged 5– 17□years: summary of the evidence. International Journal of Behavioral Nutrition and Physical Activity. 2020;17(1):141.

29. Shayo FK. Co-occurrence of risk factors for non-communicable diseases among in-school adolescents in Tanzania: an example of a low-income setting of sub-Saharan Africa for adolescence health policy actions. BMC public health. 2019;19(1):972.

30. Vyas S, Kumaranayake L. Constructing socio-economic status indices: how to use principal components analysis. Health policy and planning. 2006;21(6):459–68.

31. Lex A, Gehlenborg N, Strobelt H, Vuillemot R, Pfister H. UpSet: Visualization of Intersecting Sets. IEEE Trans Vis Comput Graph. 2014;20(12):1983–92.

32. Hagenaars JA, McCutcheon AL. Applied Latent Class Analysis. Cambridge: Cambridge University Press; 2002.

33. Harvey G. Multilevel Statistical Models, 4th Edition. West Sussex: John Wiley & Sons, Ld; 2010.

34. Akseer N, Mehta S, Wigle J, Chera R, Brickman ZJ, Al-Gashm S, et al. Non-communicable diseases among adolescents: current status, determinants, interventions and policies. BMC public health. 2020;20(1):1908.

35. Goodman SN. Toward Evidence-Based Medical Statistics. 2: The Bayes Factor. Annals of Internal Medicine. 1999;130(12):1005–13.

36. Bürkner P-C. brms: An R Package for Bayesian Multilevel Models Using Stan. 2017. 2017;80(1):J Journal of Statistical Software.

37. Linzer DA, Lewis JB. poLCA: An R Package for Polytomous Variable Latent Class Analysis. Journal of Statistical Software; Vol 1, Issue 10 (2011). 2011.

38. Alamian A, Paradis G. Clustering of chronic disease behavioral risk factors in Canadian children and adolescents. Preventive medicine. 2009;48(5):493–9.

39. Leech RM, McNaughton SA, Timperio A. The clustering of diet, physical activity and sedentary behavior in children and adolescents: a review. Int J Behav Nutr Phys Act. 2014;11:4-.

40. Matias TS, Silva KS, Silva JAd, Mello GTd, Salmon J. Clustering of diet, physical activity and sedentary behavior among Brazilian adolescents in the national school - based health survey (PeNSE 2015). BMC public health. 2018;18(1):1283.

41. Loef M, Walach H. The combined effects of healthy lifestyle behaviors on all cause mortality: a systematic review and meta-analysis. Preventive medicine. 2012;55(3):163–70.

42. Uddin R, Khan A. Sedentary behaviour is associated with overweight and obesity among adolescents: evidence from a population-based study. Acta Paediatrica. 2019;108(8):1545–6.

43. Carson V, Hunter S, Kuzik N, Gray CE, Poitras VJ, Chaput JP, et al. Systematic review of sedentary behaviour and health indicators in school-aged children and youth: an update. Applied physiology, nutrition, and metabolism = Physiologie appliquee, nutrition et metabolisme. 2016;41(6 Suppl 3):S240–65.

44. Elliott MR, Shope JT, Raghunathan TE, Waller PF. Gender differences among young drivers in the association between high-risk driving and substance use/environmental influences. Journal of studies on alcohol. 2006;67(2):252–60.

45. Nolen-Hoeksema S. Sex differences in unipolar depression: evidence and theory. Psychological bulletin. 1987;101(2):259–82.

46. Nolen-Hoeksema S, Girgus JS. The emergence of gender differences in depression during adolescence. Psychological bulletin. 1994;115(3):424–43.

47. Dumont M, Provost MA. Resilience in Adolescents: Protective Role of Social Support, Coping Strategies, Self-Esteem, and Social Activities on Experience of Stress and Depression. Journal of Youth and Adolescence. 1999;28(3):343–63.

48. Choudhry S, McClinton-Powell L, Solomon M, Davis D, Lipton R, Darukhanavala A, et al. Power-up: a collaborative after-school program to prevent obesity in African American children. Progress in community health partnerships : research, education, and action. 2011;5(4):363–73.

49. Foster GD, Sherman S, Borradaile KE, Grundy KM, Vander Veur SS, Nachmani J, et al. A policy-based school intervention to prevent overweight and obesity. Pediatrics. 2008;121(4):e794–802.

50. Lobstein T, Baur L, Uauy R. Obesity in children and young people: a crisis in public health. Obesity Reviews. 2004;5(1):4–85.

51. Fung C, Kuhle S, Lu C, Purcell M, Schwartz M, Storey K, et al. From “best practice” to “next practice”: the effectiveness of school-based health promotion in improving healthy eating and physical activity and preventing childhood obesity. Int J Behav Nutr Phys Act. 2012;9:27.

52. Faught EL, Ekwaru JP, Gleddie D, Storey KE, Asbridge M, Veugelers PJ. The combined impact of diet, physical activity, sleep and screen time on academic achievement: a prospective study of elementary school students in Nova Scotia, Canada. Int J Behav Nutr Phys Act. 2017;14(1):29.

53. Veugelers PJ, Fitzgerald AL. Effectiveness of school programs in preventing childhood obesity: a multilevel comparison. Am J Public Health. 2005;95(3):432–5.

54. Langford R, Bonell C, Jones H, Pouliou T, Murphy S, Waters E, et al. The World Health Organization’s Health Promoting Schools framework: a Cochrane systematic review and meta-analysis. BMC public health. 2015;15(1):130.

55. Ministry of Domestic Affairs of Vietnam, UNFPA. National report on Vietnames youth. Hanoi, Vietnam 2015.

