## Supplemental material for "Clustering Lifestyle Risk Behaviors among Vietnamese Adolescents and Roles of School: A Bayesian Multilevel Analysis of Global School-Based Student Health Survey 2019"

### Supplemental materials S1

**Supplemental table 1: Literature review of studies on risk factors clustering among adolescents**

| **No.** | **Name** | **Author (year)** | **Country** | **Data source** | **Number of behaviors** | **Name of behaviors** | **Analytic approach for clustering** |
| --- | --- | --- | --- | --- | --- | --- | --- |
| 1 | The clustering of risk behaviours in adolescence and health consequences in middle age [1] | Akasaki M (2019) | England, Wales and Scotland | British Cohort Study | 7 | Smoking, multiple binge drinking, drug use, early sexual intercourse, unprotected sex, physical inactivity, involvement in fights and delinquency | Latent class analysis |
| 2 | Clustering of chronic disease behavioral risk factors in Canadian children and adolescents [2] | Alamian A (2009) | Canada | National Longitudinal Survey of Children and Youth (Cycle 4) | 5 | Physical inactivity, sedentary behavior, ever smoking, ever drinking, high BMI | Ratios of observed (O) and expected (E) |
| 3 | Clustering of oral and general health risk behaviors among adolescents [3] | Austregésilo SC (2019) | Korea | Korea Youth Risk Behavior Web-based Survey | 4 | Frequent smoking, drunkenness, regular breakfast, regular exercise | Treating the effects of risk behaviors equally |
| 4 | Association between Clustering of Lifestyle Behaviors and Health-Related Physical Fitness in Youth: The UP&DOWN Study [4] | Cabanas-Sánchez V (2018) | Spain | UP&DOWN study | 5 | Screen time, nonscreen sedentary time, moderate-to-vigorous physical activity, mediterranean diet quality, sleep time | Hierarchical clustering and k-means clustering |
| 5 | Prevalence of cardiometabolic risk factor clustering and body mass index in adolescents [5] | Camhi SM (2011) | The US | National Health and Nutrition Examination Survey | 4 | High level of triglycerides, high-density lipoprotein cholesterol, systolic/diastolic blood pressure, fasting glucose | Treating the effects of risk behaviors equally |
| 6 | Clustering of lifestyle factors and the relationship with depressive symptoms among adolescents in Northeastern China [6] | Cao R (2020) | China | A cross-sectional study in Jilin Province, China | 3 | Screen time, physical activity, sleep duration | Two-step cluster analysis |
| 7 | Gender differences in factors influencing smoking, drinking, and their co-occurrence among adolescents in South Korea [7] | Chun J (2013) | Korea | Korean Youth Panel Study (KYPS) | 2 | Smoking, drinking | Treating the effects of risk behaviors equally |
| 8 | Prevalence and factors associated with the co-occurrence of health risk behaviors in adolescents [8] | da Silva Brito AL (2015) | Brazil | Lifestyle and health risk behaviors in adolescents: from prevalence study to intervention | 5 | Low level of physical activity, sedentary behavior, occasional consumption of fruits and vegetables, alcohol consumption, smoking | Treating the effects of risk behaviors equally |
| 9 | Clustering of chronic diseases risk factors among adolescents: a quasi-experimental study in Sousse, Tunisia [9] | Dendana E (2017) | Tunisia | A quasi-experimental school-based intervention in Sousse, Tunisia | 4 | Smoking, sedentary behavior, low fruit and vegetable intake, obesity | Treating the effects of risk behaviors equally |
| 10 | Prevalence, clustering and sociodemographic distributions of non-communicable disease risk factors in Nepalese adolescents: secondary analysis of a nationwide school survey [10] | Dhungana RR (2019) | Nepal | The Global School-based Student Health Survey | 5 | Smoking, alcohol consumption, insufficient fruit and vegetable, insufficient physical activity, overweight and obesity | Treating the effects of risk behaviors equally |
| 11 | Clustering of Six Key Risk Behaviors for Chronic Disease among Adolescent Females [11] | Gardner LA (2020) | Australia | A cross-sectional study in seven Australian independent secondary schools | 6 | Physical inactivity, poor diet, recreational screen time, inadequate sleep, alcohol use, and smoking | Latent class analysis |
| 12 | Perceived peer norms, health risk behaviors, and clustering of risk behaviors among Palestinian youth [12] | Glick P (2018) | Palestine | A cross-sectional study in West Bank and East Jerusalem | 4 | Smoking, alcohol use, drug use, sexual intercourse | Treating the effects of risk behaviors equally |
| 13 | Risky behaviors in late adolescence: co-occurrence, predictors, and consequences [13] | Hair EC (2009) | The US | National Longitudinal Survey of Youth | 6 | No exercise, delinquency, smoke cigarettes, drug use, binge drinking, unsafe sex | Latent class analysis |
| 14 | Clustering of Health Behaviors and Cardiorespiratory Fitness Among U.S. Adolescents [14] | Hartz J (2018) | The US | National Health and Nutrition Examination Survey | 3 | Dietary intake and quality, physical activity, sedentary time | K-mean clustering |
| 15 | Clustering patterns of oral and general health-risk behaviors in Brazilian adolescents: Findings from a national survey [15] | Jordão LMR (2018) | Brazil | Brazilian National School-based Student Health Survey | 17 | Eating while watching TV or studying, insufficient physical activity, low fruit intake, high intake of sweets, no dental visits (last 12 mo), high intake of soft drinks, high intake of biscuits, current drinking, skipping breakfast, physical fighting, no car seatbelt use compliance, less frequent tooth brushing, no motorcycle helmet use compliance, unprotected sex, current smoking, no hand washing before meals, current illegal drug use | Hierarchical cluster analysis |
| 16 | lustering of risk-related modifiable behaviours and their association with overweight and obesity among a large sample of youth in the COMPASS study [16] | Laxer RE (2017) | Canada | COMPASS | 15 | Physical activity, dietary behaviors, sedentary behaviors, substance use behaviors | Latent class analysis |
| 17 | An examination of the co-occurrence of modifiable risk factors associated with chronic disease among youth in the COMPASS study [17] | Leatherdale ST (2015) | Canada | COMPASS | 7 | Smoking, marijuana use, binge drinking, overweight/obese, physically Inactive, highly sedentary, inadequate fruit & vegetable consumption | Treating the effects of risk behaviors equally |
| 18 | Socioeconomic Disparities in Health Risk Behavior Clusterings Among Korean Adolescents [18] | Lee B (2018) | Korea | Korean Youth Risk Behavior Survey | 3 | Cigarette smoking, drinking, unprotected sex | Treating the effects of risk behaviors equally |
| 19 | Dietary habits and physical activity: Results from cluster analysis and market basket analysis [19] | Liew HP (2018) | The US | The 2011 Healthy School Program (HSP) Evaluation | 2 | Quality of foods, dietary habits, physical activity | K-mean clustering |
| 20 | Cluster of risk and protective factors for obesity among Brazilian adolescents [20] | Maia EG (2018) | Brazil | The Brazilian National School Health Survey of 2012 | 4 | Dietary intake, eating behavior, physical activity, sedentary behaviors | K-mean clustering |
| 21 | Co-occurrence of Health Risk Behaviors Among Brazilian Adolescent Victims of Family Violence [21] | Marques ES (2018) | Brazil | The National School Health Survey | 4 | Fights, substance use (alcohol, drugs, and tobacco), not wearing seat belts and helmets, inadequate food consumption | Treating the effects of risk behaviors equally |
| 22 | Clusters of health behaviours and their relation to body mass index among adolescents in Northern Finland [22] | Marttila-Tornio K (2020) | Finland | The prospective population-based Northern Finland Birth Cohort 1986 | 7 | Physical activity, screen time, cigarette smoking, alcohol use, sugary foods intake, fast food intake, fruit, vegetable & berries intake | K-mean clustering |
| 23 | Clustering of diet, physical activity and sedentary behavior among Brazilian adolescents in the national school - based health survey (PeNSE 2015) [23] | Matias TS (2018) | Brazil | The 2015 National School-Based Health Survey | 3 | Diet, physical activity, sedentary behavior | K-mean clustering |
| 24 | Clustering of multiple energy balance related behaviors is associated with body fat composition indicators in adolescents: Results from the HELENA and ELANA studies [24] | Moreira NF (2018) | 8 European countries & Brazil | Healthy Lifestyle in Europe by Nutrition in Adolescents the Adolescent Nutritional Assessment Longitudinal Study | 4 | Television watching, moderate and vigorous physical activity, consumption of fruits and vegetables, consumption of sugar-sweetened beverages | K-mean clustering |
| 25 | Adolescent BMI trajectories with clusters of physical activity and sedentary behaviour: an exploratory analysis [25] | Nesbit KC (2016) | The US | The National Institute of Child Health and Human Development Study of Early Child Care and Youth Development | 3 | Physical activity (monitor; child report/parent report), TV/video watching time, PC recreational use | Treating the effects of risk behaviors equally |
| 26 | Clustering of energy balance-related behaviours, sleep, and overweight among Finnish adolescents [26] | Nuutinen T (2017) | Finland | The cross-national Health Behaviour in School-aged Children (HBSC) study | 6 | Sleep duration, discrepancy and quality, physical activity, screen time, junk food, fruit and vegetable intake | K-mean clustering |
| 27 | Gender and Health Behavior Clustering among U.S. Young Adults [27] | Olson JS (2017) | The US | Add Health | 10 | Binge drink, cigarette smoke, other tobacco use, physical activity, marijuana use, doctor visit, dentist visit, fast food consumption, abused prescription drugs, illegal drug use | Latent class analysis |
| 28 | Clustering of oral and general health risk behaviors in Korean adolescents: a national representative sample [28] | Park YD (2010) | Korea | The Korea Youth Risk Behavior Web-based Survey | 6 | Frequent tooth brushing, preventive dental care, tobacco use, alcohol misuse, eating, physical activity | Treating the effects of risk behaviors equally |
| 29 | Cluster patterns of behavioural risk factors among children: Longitudinal associations with adult cardio-metabolic risk factors [29] | Patterson KAE (2019) | Australia | The Australian Schools Health and Fitness Survey | 5 | Smoking status, alcohol consumption, physical activity, breakfast consumption, psychological well-being | TwoStep cluster analysis |
| 30 | Lifestyle Clusters in School-Aged Youth and Longitudinal Associations with Fatness: The UP&DOWN Study [30] | Sánchez-Oliva D (2018) | Spain | The UP&DOWN study | 5 | Screen time, sedentary time, moderate to vigorous physical activity, healthy diet, obesity | K-mean clustering |
| 31 | Co-occurrence of risk factors for non-communicable diseases among in-school adolescents in Tanzania: an example of a low-income setting of sub-Saharan Africa for adolescence health policy actions [31] | Shayo FK (2019) | Tanzania | The 2014 Tanzania Global School-based Student Health Survey | 5 | An unhealthy diet, physical inactivity, tobacco use, excessive alcohol use, suicide attempt | Treating the effects of risk behaviors equally |
| 32 | Gender differences in the clustering patterns of risk behaviours associated with non-communicable diseases in Brazilian adolescents [32] | Silva KS (2014) | Brazil | Lifestyle and risk behaviours of young people in Santa Catarina State, Southern Brazil—COMPAC project | 4 | Excessive screen-time, insufficient moderate to vigorous physical activity, low fruit/vegetable intake, consumption of alcohol | Ratios of observed (O) and expected (E) |
| 33 | A cluster-analytic approach towards multidimensional health-related behaviors in adolescents: the MoMo-Study [33] | Spengler S (2012) | Germany | the German Health Interview and Examination Survey for Children and Adolescents (KiGGS)  the “Motorik-Modul” (MoMo) | 3 | Food consumption, physical activity, electronic media use | K-mean clustering |
| 34 | Clustering of health risk behaviors among adolescents in Kilifi, Kenya, a rural Sub-Saharan African setting [34] | Ssewanyana D (2020) | Kenya | The Kilifi Health and Demographic Surveillance System | 5 | Injury and violence-related behavior, substance use, hygiene behavior, physical activity, dietary behavior | Latent class analysis |
| 35 | Do sedentary behavior and physical activity spatially cluster? Analysis of a population-based sample of Boston adolescents [35] | Tamura K (2018) | The US | The 2008 Boston Youth Survey Geospatial Dataset | 4 | TV watching, video games, total screen time, 20min/day of physical activity | Spatial clustering |
| 36 | Clustering of lifestyle risk behaviours and its determinants among school-going adolescents in a middle-income country: a cross-sectional study [36] | Teh C H (2019) | Malaysia | The Malaysian Adolescent Health Risk Behaviour study | 5 | Smoking, alcohol consumption, physical inactivity, sedentary behavior, low fruit/vegetable intake | Ratios of observed (O) and expected (E) |
| 37 | A cluster analysis of physical activity and sedentary behavior patterns in middle school girls [37] | Trilk JL (2012) | The US | The Trial of Activity for Adolescent Girls | 6 | Educational sedentary, sports and play, past year organized sports/teams/lessons, active transportation and chores, electronic media, sleep | K-mean clustering |
| 38 | Clustering of lifestyle risk factors for non-communicable diseases in 304,779 adolescents from 89 countries: A global perspective [38] | Uddin R (2020) | 89 countries | The Global School-based Student Health Survey | 6 | Low fruit-vegetable, physical inactivity, high sedentary behavior, overweight/obesity, currently drink alcohol, currently smoke | Ratios of observed (O) and expected (E) |
| 39 | Association between Cluster of Lifestyle Behaviors and HOMA-IR among Adolescents: ABCD Growth Study [39] | Werneck AO (2018) | Brazil | Analysis of Behaviors of Children During Growth | 4 | Skipping breakfast, poor sleep quality, no sport participation, high TV viewing | Treating the effects of risk behaviors equally |

### Supplemental materials S2

**Supplemental Table 2: Variable definition**

| **Variables** | **GSHS 2019 questionnaire** | **Definition used in this paper** |
| --- | --- | --- |
| **Lifestyle risk behaviors** | | |
| Smoking | During the past 30 days, on how many days did you smoke cigarettes? | Students who smoked at least 1 day in the past 30 days  *Yes vs.No* |
| Alcohol consumption | During the past 30 days, on how many days did you have at least one drink containing alcohol? | Students who drank at least one alcoholic beverage in the past 30 days  *Yes vs.No* |
| Physical inactivity | During the past 7 days, on how many days were you physically active for a total of at least 60 minutes per day? | Students who did not spend at least 60 min per day on moderate-to-vigorous intensity physical activities, as across the week  *Yes vs.No* |
| Sedentary behavior | How much time do you spend during a typical or usual day sitting and watching television, playing computer games, relaxing with Ipad, mobile phone, talking with friends, or doing other sitting activities, such as reading books or using Facebook? | Students who spent at least 3 hours a day doing these activities  *Yes vs.No* |
| Low fruit/vegetable intake | - During the past 30 days, on average how many times per day did you usually eat fruit, such as a banana, apple, orange, guava, rambutan, watermelon, papaya, or mango, etc.  - During the past 30 days, on average, how many times per day did you usually eat vegetables, such as morning glory, cabbage, or mustard green, etc.? | Students who did not consume both fruits and vegetables at least two times per day.  *Yes vs.No* |
| Unhealthy diet | - During the past 30 days, on average how many times per day did you usually drink carbonated soft drinks, such as Coca Cola, Pepsi, or Fanta? (Do not include diet soft drinks, such as Coke Zero or Diet Coke.)  - During the past 7 days, on how many days did you eat food at least once a day from a fast food restaurant, such as KFC, Lotteria, Circle K etc.? | Students who drank carbonated soft drinks at least one time per day or ate fast-food at least one day a week.  *Yes vs.No* |
| **Student-level variables** | | |
| Sex |  | *Male vs. Female* |
| Age |  | *13 – 17* |
| BMI-Z score | *The BMI Z score was calculated based on the WHO 2007 reference for children aged 5-19 years with the Stata macro package that is available at* [*https://www.who.int/growthref/tools/readme_stata.pdf?ua=1*](https://www.who.int/growthref/tools/readme_stata.pdf?ua=1)*.* | |
| Place of residence |  | *Rural vs. Urban* |
| Living with | Do you live with both of your parents? | Students who are living with father and/or mother  *Yes vs.No* |
| Parents monitoring | - During the past 30 days, how often did your parents or guardians check to see if your homework was done?  - During the past 30 days, how often did your parents or guardians really know what you were doing with your free time? (*1 “Never” to 5 “Always”*) | Total score, ranging from 1 to 10 |
| Number of close friends | How many close friends do you have? | *0, 1, 2, 3 or more* |
| Loneliness | During the past 12 months, how often have you felt lonely? (*1 “Never” to 5 “Always”*) | Students who most of the time/always felt lonely during the past 12 months  *Yes vs. No* |
| Worrying | During the past 12 months, how often have you been so worried about something that you could not eat or did not feel hungry? (*1 “Never” to 5 “Always”*) | Students who most of the time/always felt worry during the past 12 months  *Yes vs. No* |
| Truancy | During the past 30 days, on how many days did you miss classes or school without permission? | Students who missed any days without permission during the past 30 days  *Yes vs. No* |
| **School health promotion programs** | | |
| During the last school year, were you taught in any of your classes the problems associated with using drugs, such as drug addiction and crime? | | Aggregated at the school level (%) |
| During the last school year, were you taught in any of your classes the problems associated with drinking alcohol? | | Aggregated at the school level (%) |
| During the last school year, were you taught about the benefits of physical activity? | | Aggregated at the school level (%) |
| During the last school year, were you taught in any of your classes signs of depression and suicidal behavior? | | Aggregated at the school level (%) |
| During the last school year, were you taught in any of your classes the benefits of eating more fruits and vegetables? | | Aggregated at the school level (%) |

### Supplemental materials S3:

**Latent class analysis (LCA) of lifestyle risk behaviors clustering**

We used latent class analysis to identify homogeneous unobservable subgroups of lifestyle risk behaviors. The final number of classes were determined based on the empirical evidence and interpretability of the class memberships. Since two latent classes was the most interpretable, we decided to choose two latent classes as the outcome for the clustering of lifestyle risk behaviors modeling.

Predicted mean

|  | Class 1 | Class 2 |
| --- | --- | --- |
| Smoking | 0.0021 | 0.1247 |
| Drinking | 0.1057 | 0.706 |
| Physical inactivity | 0.8562 | 0.8324 |
| Sedentary behavior | 0.4255 | 0.6777 |
| Low fruit/vegetable intake | 0.2946 | 0.3908 |
| Unhealthy diet | 0.6199 | 0.7823 |

**Supplemental Figure 1: Distribution of lifestyle risk behaviors for two classes of LCA**

**Supplemental Figure 2: Distribution of lifestyle risk behaviors for three classes of LCA**

Predicted mean

|  | Class 1 | Class 2 | Class 3 |
| --- | --- | --- | --- |
| Smoking | 0.0084 | 0.126 | 0.127285 |
| Drinking | 0.1463 | 0.7395 | 0.083 |
| Physical inactivity | 0.7517 | 0.8358 | 0.9126 |
| Sedentary behavior | 0.2108 | 0.7009 | 0.5417 |
| Low fruit/vegetable intake | 0.067 | 0.3977 | 0.4214 |
| Unhealthy diet | 0.6543 | 0.7804 | 0.6055 |

Predicted mean

|  | Class 1 | Class 2 | Class 3 | Class 4 |
| --- | --- | --- | --- | --- |
| Smoking | 0 | 0.0083 | 0.0457 | 0.1276 |
| Drinking | 0.1016 | 0.0964 | 0.2533 | 0.7755 |
| Physical inactivity | 0.896 | 0.898 | 0.3947 | 0.8521 |
| Sedentary behavior | 0.4175 | 0.5241 | 0.1969 | 0.7074 |
| Low fruit/vegetable intake | 0.0021 | 0.9829 | 0.0225 | 0.3988 |
| Unhealthy diet | 0.6419 | 0.5889 | 0.6228 | 0.7926 |

**Supplemental Figure 3: Distribution of lifestyle risk behaviors for four classes of LCA**

Predicted mean

|  | Class 1 | Class 2 | Class 3 | Class 4 | Class 5 |
| --- | --- | --- | --- | --- | --- |
| Smoking | 0.135 | 0 | 0.1916 | 0 | 0.0148 |
| Drinking | 0.6485 | 0.069 | 0.7768 | 0.2587 | 0.1515 |
| Physical inactivity | 0.643 | 0.8176 | 0.8508 | 0.925 | 0.9032 |
| Sedentary behavior | 0.3992 | 0.2398 | 0.7353 | 0.7396 | 0.5378 |
| Low fruit/vegetable intake | 0 | 0.116 | 0.5607 | 0.2377 | 0.9914 |
| Unhealthy diet | 0.7219 | 0.6191 | 0.8702 | 0.6822 | 0.5488 |

**Supplemental Figure 4: Distribution of lifestyle risk behaviors for five classes of LCA**

### Supplemental materials S4

**Conceptual framework for covariates of** **effects of school health promotion programs on lifestyle risk behaviors**

We developed an evidence-based conceptual framework to explain potential associations between school health promotion programs and lifestyle risks behaviors co-occurrence which is fit for Vietnam’s setting. This conceptual framework was based on the conceptual framework of risk and protective factors contribute to NCDs among adolescents developed by Akseer et al [40].

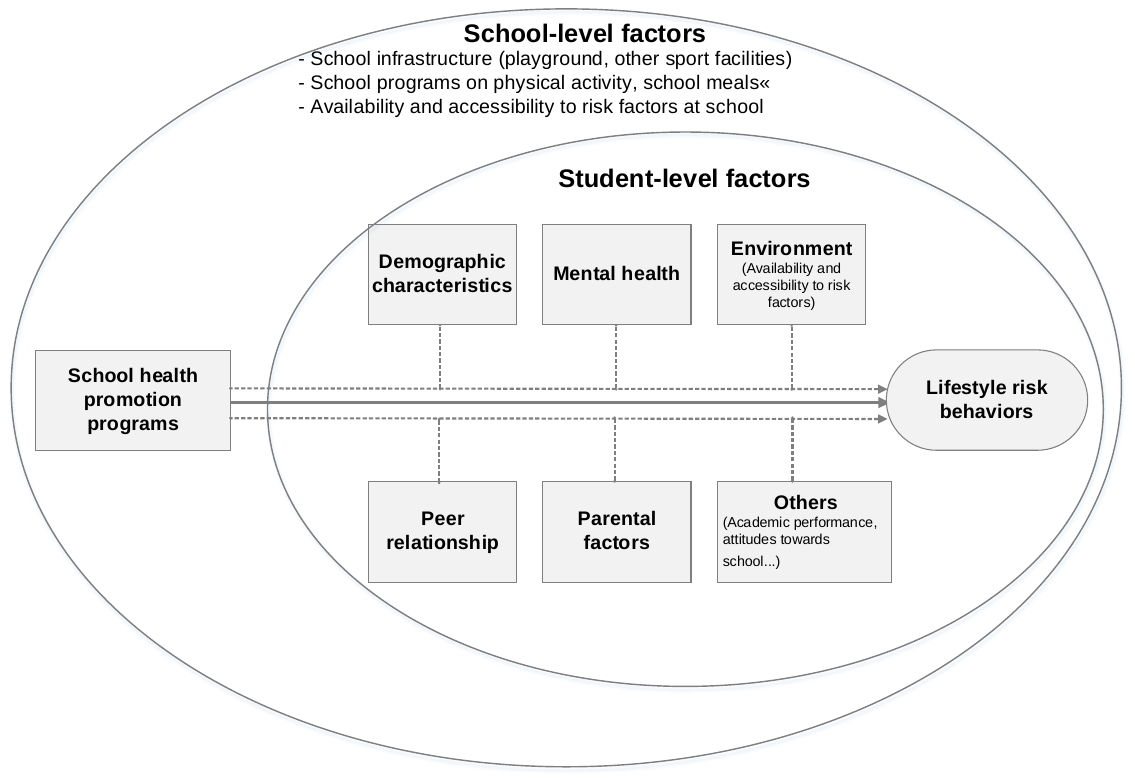

**Supplemental Figure 5:** **Conceptual framework for covariates of effects of school health promotion programs on lifestyle risk behaviors**

**Bayesian models for sensitivity analysis of effects of school health promotion programs on lifestyle risk behaviors**

***Prior construction***

We conducted sensitivity analyses to test whether the school effects remain consistent across different priors. We constructed two priors to reflect two degrees of belief that the school health promotion programs had either no impact (*equivocal prior*) or positively impact (*optimistic prior*) on lifestyle risk behaviors of students. We assumed that prior distributions of school effects were the normal distribution (i.e., *β_1_* ~ Normal(μ, σ^2^)).

In the *equivocal prior*, we assumed that the school health promotion programs did not affect lifestyle risk behaviors (i.e., *β_1_* = log(OR) = 0), the school effects can be either positive or negative with equal chance. The probability that school health promotion programs increases/decreases the odds of high level of lifestyle risk behaviors by 50% was small (i.e., 2·5%), or P[*β_1_* = log(OR) > 0·41] = 0·025. Thus we translated this prior to a normal distribution with μ = 0 and σ = 0·41/Z_1-0·025_ = 0·207.

In the *optimistic prior*, we hypothesized that the school health promotion programs could decrease the odds of high level of lifestyle risk behaviors by 50% with the same SD as in *equivocal prior.* Therefore, in this scenario, the prior was a normal distribution with μ = log(0·5) = -0·693 and σ = 0·207. These two priors were summarized as:

Equivocal prior: *β_1_* ~ Normal (0, 0·207^2^)

Optimistic prior: *β_1_* ~ Normal(-0.693, 0·207^2^)

**Supplemental Table 3: Bayesian models with different priors of effects of school health promotion programs on lifestyle risk behaviors among male adolescents in Vietnam**

|  | **Model 1** | | **Model 2** | | **Model 3^§^** | | **Probability (BF)** | | |
| --- | --- | --- | --- | --- | --- | --- | --- | --- | --- |
|  | **OR** | **95% HDI** | **OR** | **95% HDI** | **OR** | **95% HDI** | **OR < 1** | **OR < 0·9** | **OR < 0·8** |
| **Fixed part** |  |  |  |  |  |  |  |  |  |
| **Intercept** | 0·30 | 0·25 – 0·35 | 0·35 | 0·26 – 0·47 | 0·24 | 0·14 – 0·43 |  |  |  |
| **Scenario 1: Vage prior** |  |  |  |  |  |  |  |  |  |
| **School quality proxy** *(Ref: 1^st^ tertile)* |  |  |  |  |  |  |  |  |  |
| 2nd tertile |  |  | 1·08 | 0·73 – 1·62 | 0·98 | 0·70 – 1·38 | 54·6 (1·2) | 31·0 (0·45) | 11·6 (0·13) |
| 3rd tertile |  |  | **0·58** | **0·39 – 0·88** | **0·67** | **0·46 – 0·93** | 98·8 (84·4) | 95·7 (22·3) | 85·3 (5·8) |
| **Scenario 2: Equivocal prior** |  |  |  |  |  |  |  |  |  |
| **School quality proxy** *(Ref: 1^st^ tertile)* |  |  |  |  |  |  |  |  |  |
| 2nd tertile |  |  | 1·12 | 0·85 – 1·47 | 1·03 | 0·81 – 1·33 | 39·2 (0·64) | 13·3 (0·15) | 1·8 (0·02) |
| 3rd tertile |  |  | **0·74** | **0·56 – 0·98** | 0·78 | 0·60 – 1·01 | 96·8 (30·3) | 85·4 (5·8) | 56·4 (1·29) |
| **Scenario 3: Optimistic prior** |  |  |  |  |  |  |  |  |  |
| **School quality proxy** *(Ref: 1^st^ tertile)* |  |  |  |  |  |  |  |  |  |
| 2nd tertile |  |  | **0·74** | **0·55 – 0·97** | **0·73** | **0·57 – 0·95** | 99·3 (133·6) | 95·0 (18·8) | 76·4 (3·2) |
| 3rd tertile |  |  | **0·49** | **0·37 – 0·64** | **0·55** | **0·43 – 0·72** | 100 (>10000) | 100 (>10000) | 99·8 (537·5) |

BF: Bayes factor; HDI: Highest density interval; OR: Odds ratio

**^§^**Model 3 was adjusted for age, body mass index, place of residence, living with mother/father, parental monitoring, number of close friend, loneliness, worrying, and truancy

**Supplemental Table 4. Bayesian models with different priors of relationship between school health promotion programs and lifestyle risk behaviors among female adolescents in Vietnam**

|  | **Model 1** | | **Model 2** | | **Model 3^§^** | | **Probability (BF)** | | |
| --- | --- | --- | --- | --- | --- | --- | --- | --- | --- |
|  | **OR** | **95% HDI** | **OR** | **95% HDI** | **OR** | **95% HDI** | **OR < 1** | **OR < 0·9** | **OR < 0·8** |
| **Fixed part** |  |  |  |  |  |  |  |  |  |
| **Intercept** | 0·20 | 0·17 – 0·24 | 0·23 | 0·17 – 0·30 | 0·13 | 0·08 – 0·23 |  |  |  |
| **Scenario 1: Vage prior** |  |  |  |  |  |  |  |  |  |
| **School quality proxy** *(Ref: 1^st^ tertile)* |  |  |  |  |  |  |  |  |  |
| 2nd tertile |  |  | 1·18 | 0·77 – 1·78 | 1·10 | 0·77 – 1·59 | 29·6 (0·42) | 13·1 (0·15) | 4·0 (0·04) |
| 3rd tertile |  |  | **0·64** | **0·42 – 0·94** | **0·69** | **0·46 – 0·98** | 97·4 (37·1) | 91·5 (10·7) | 77·3 (3·4) |
| **Scenario 2: Equivocal prior** |  |  |  |  |  |  |  |  |  |
| **School quality proxy** *(Ref: 1^st^ tertile)* |  |  |  |  |  |  |  |  |  |
| 2nd tertile |  |  | 1·16 | 0·88 – 1·54 | 1·11 | 0·86 – 1·45 | 21·3 (0·27) | 5·0 (0·06) | 1·1 (0·01) |
| 3rd tertile |  |  | 0·77 | 0·58 – 1·04 | 0·80 | 0·62 – 1·05 | 94·8 (18·1) | 80·4 (4·1) | 49·7 (1·0) |
| **Scenario 3: Optimistic prior** |  |  |  |  |  |  |  |  |  |
| **School quality proxy** *(Ref: 1^st^ tertile)* |  |  |  |  |  |  |  |  |  |
| 2nd tertile |  |  | **0·75** | **0·55 – 1·00** | **0·75** | **0·57 – 0·99** | 98·2 (55·2) | 90·1 (9·1) | 66·4 (2·0) |
| 3rd tertile |  |  | **0·50** | **0·37 – 0·66** | **0·54** | **0·41 – 0·70** | 100 (>10000) | 100 (>10000) | 99·8 (499) |

BF: Bayes factor; HDI: Highest density interval; OR: Odds ratio

**^§^**Model 3 was adjusted for age, body mass index, place of residence, living with mother/father, parental monitoring, number of close friend, loneliness, worrying, and truancy

### References

1.	Akasaki M, Ploubidis GB, Dodgeon B, Bonell CP. The clustering of risk behaviours in adolescence and health consequences in middle age. Journal of adolescence. 2019;77:188-97.

2.	Alamian A, Paradis G. Clustering of chronic disease behavioral risk factors in Canadian children and adolescents. Preventive medicine. 2009;48(5):493-9.

3.	Austregésilo SC, de Goes PSA, de Sena Júnior MR, Pazos CTC. Clustering of oral and general health risk behaviors among adolescents. Preventive medicine reports. 2019;15:100936.

4.	Cabanas-Sánchez V, Martínez-Gómez D, Izquierdo-Gómez R, Segura-Jiménez V, Castro-Piñero J, Veiga OL. Association between Clustering of Lifestyle Behaviors and Health-Related Physical Fitness in Youth: The UP&DOWN Study. The Journal of pediatrics. 2018;199:41-8.e1.

5.	Camhi SM, Katzmarzyk PT. Prevalence of cardiometabolic risk factor clustering and body mass index in adolescents. The Journal of pediatrics. 2011;159(2):303-7.

6.	Cao R, Gao T, Hu Y, Qin Z, Ren H, Liang L, et al. Clustering of lifestyle factors and the relationship with depressive symptoms among adolescents in Northeastern China. Journal of affective disorders. 2020;274:704-10.

7.	Chun J, Chung IJ. Gender differences in factors influencing smoking, drinking, and their co-occurrence among adolescents in South Korea. Nicotine & tobacco research : official journal of the Society for Research on Nicotine and Tobacco. 2013;15(2):542-51.

8.	da Silva Brito AL, Hardman CM, de Barros MV. [Prevalence and factors associated with the co-occurrence of health risk behaviors in adolescents]. Revista paulista de pediatria : orgao oficial da Sociedade de Pediatria de Sao Paulo. 2015;33(4):423-30.

9.	Dendana E, Ghammem R, Sahli J, Maatoug J, Fredj SB, Harrabi I, et al. Clustering of chronic diseases risk factors among adolescents: a quasi-experimental study in Sousse, Tunisia. International journal of adolescent medicine and health. 2017;31(4).

10.	Dhungana RR, Bista B, Pandey AR, de Courten M. Prevalence, clustering and sociodemographic distributions of non-communicable disease risk factors in Nepalese adolescents: secondary analysis of a nationwide school survey. BMJ open. 2019;9(5):e028263.

11.	Gardner LA, Champion KE, Parmenter B, Grummitt L, Chapman C, Sunderland M, et al. Clustering of Six Key Risk Behaviors for Chronic Disease among Adolescent Females. International journal of environmental research and public health. 2020;17(19).

12.	Glick P, Khammash U, Shaheen M, Brown R, Goutam P, Karam R, et al. Perceived peer norms, health risk behaviors, and clustering of risk behaviors among Palestinian youth. PloS one. 2018;13(6):e0198435.

13.	Hair EC, Park MJ, Ling TJ, Moore KA. Risky behaviors in late adolescence: co-occurrence, predictors, and consequences. The Journal of adolescent health : official publication of the Society for Adolescent Medicine. 2009;45(3):253-61.

14.	Hartz J, Yingling L, Ayers C, Adu-Brimpong J, Rivers J, Ahuja C, et al. Clustering of Health Behaviors and Cardiorespiratory Fitness Among U.S. Adolescents. The Journal of adolescent health : official publication of the Society for Adolescent Medicine. 2018;62(5):583-90.

15.	Jordão LMR, Malta DC, Freire M. Clustering patterns of oral and general health-risk behaviours in Brazilian adolescents: Findings from a national survey. Community dentistry and oral epidemiology. 2018;46(2):194-202.

16.	Laxer RE, Brownson RC, Dubin JA, Cooke M, Chaurasia A, Leatherdale ST. Clustering of risk-related modifiable behaviours and their association with overweight and obesity among a large sample of youth in the COMPASS study. BMC public health. 2017;17(1):102.

17.	Leatherdale ST. An examination of the co-occurrence of modifiable risk factors associated with chronic disease among youth in the COMPASS study. Cancer causes & control : CCC. 2015;26(4):519-28.

18.	Lee B, Seo DC. Socioeconomic Disparities in Health Risk Behavior Clusterings Among Korean Adolescents. International journal of behavioral medicine. 2018;25(5):540-7.

19.	Liew HP. Dietary habits and physical activity: Results from cluster analysis and market basket analysis. Nutrition and health. 2018;24(2):83-92.

20.	Maia EG, Mendes LL, Pimenta AM, Levy RB, Claro RM. Cluster of risk and protective factors for obesity among Brazilian adolescents. International journal of public health. 2018;63(4):481-90.

21.	Marques ES, Azeredo CM, de Oliveira A. Co-occurrence of Health Risk Behaviors Among Brazilian Adolescent Victims of Family Violence. Journal of interpersonal violence. 2018:886260518786493.

22.	Marttila-Tornio K, Ruotsalainen H, Miettunen J, Männikkö N, Kääriäinen M. Clusters of health behaviours and their relation to body mass index among adolescents in Northern Finland. Scandinavian journal of caring sciences. 2020;34(3):666-74.

23.	Matias TS, Silva KS, Silva JAD, Mello GT, Salmon J. Clustering of diet, physical activity and sedentary behavior among Brazilian adolescents in the national school - based health survey (PeNSE 2015). BMC public health. 2018;18(1):1283.

24.	Moreira NF, da Veiga GV, Santaliestra-Pasías AM, Androutsos O, Cuenca-García M, de Oliveira ASD, et al. Clustering of multiple energy balance related behaviors is associated with body fat composition indicators in adolescents: Results from the HELENA and ELANA studies. Appetite. 2018;120:505-13.

25.	Nesbit KC, Low JA, Sisson SB. Adolescent BMI trajectories with clusters of physical activity and sedentary behaviour: an exploratory analysis. Obesity science & practice. 2016;2(2):115-22.

26.	Nuutinen T, Lehto E, Ray C, Roos E, Villberg J, Tynjälä J. Clustering of energy balance-related behaviours, sleep, and overweight among Finnish adolescents. International journal of public health. 2017;62(8):929-38.

27.	Olson JS, Hummer RA, Harris KM. Gender and Health Behavior Clustering among U.S. Young Adults. Biodemography and social biology. 2017;63(1):3-20.

28.	Park YD, Patton LL, Kim HY. Clustering of oral and general health risk behaviors in Korean adolescents: a national representative sample. The Journal of adolescent health : official publication of the Society for Adolescent Medicine. 2010;47(3):277-81.

29.	Patterson KAE, Ferrar K, Gall SL, Venn AJ, Blizzard L, Dwyer T, et al. Cluster patterns of behavioural risk factors among children: Longitudinal associations with adult cardio-metabolic risk factors. Preventive medicine. 2020;130:105861.

30.	Sánchez-Oliva D, Grao-Cruces A, Carbonell-Baeza A, Cabanas-Sánchez V, Veiga OL, Castro-Piñero J. Lifestyle Clusters in School-Aged Youth and Longitudinal Associations with Fatness: The UP&DOWN Study. The Journal of pediatrics. 2018;203:317-24.e1.

31.	Shayo FK. Co-occurrence of risk factors for non-communicable diseases among in-school adolescents in Tanzania: an example of a low-income setting of sub-Saharan Africa for adolescence health policy actions. BMC public health. 2019;19(1):972.

32.	Silva KS, Barbosa Filho VC, Del Duca GF, de Anselmo Peres MA, Mota J, Lopes Ada S, et al. Gender differences in the clustering patterns of risk behaviours associated with non-communicable diseases in Brazilian adolescents. Preventive medicine. 2014;65:77-81.

33.	Spengler S, Mess F, Mewes N, Mensink GB, Woll A. A cluster-analytic approach towards multidimensional health-related behaviors in adolescents: the MoMo-Study. BMC public health. 2012;12:1128.

34.	Ssewanyana D, Abubakar A, Newton C, Otiende M, Mochamah G, Nyundo C, et al. Clustering of health risk behaviors among adolescents in Kilifi, Kenya, a rural Sub-Saharan African setting. PloS one. 2020;15(11):e0242186.

35.	Tamura K, Duncan DT, Athens J, Scott M, Rienti M, Jr., Aldstadt J, et al. Do sedentary behavior and physical activity spatially cluster? Analysis of a population-based sample of Boston adolescents. GeoJournal. 2018;83(4):775-82.

36.	Teh CH, Teh MW, Lim KH, Kee CC, Sumarni MG, Heng PP, et al. Clustering of lifestyle risk behaviours and its determinants among school-going adolescents in a middle-income country: a cross-sectional study. BMC public health. 2019;19(1):1177.

37.	Trilk JL, Pate RR, Pfeiffer KA, Dowda M, Addy CL, Ribisl KM, et al. A cluster analysis of physical activity and sedentary behavior patterns in middle school girls. The Journal of adolescent health : official publication of the Society for Adolescent Medicine. 2012;51(3):292-8.

38.	Uddin R, Lee EY, Khan SR, Tremblay MS, Khan A. Clustering of lifestyle risk factors for non-communicable diseases in 304,779 adolescents from 89 countries: A global perspective. Preventive medicine. 2020;131:105955.

39.	Werneck AO, Agostinete RR, Cayres SU, Urban JB, Wigna A, Chagas LGM, et al. Association between Cluster of Lifestyle Behaviors and HOMA-IR among Adolescents: ABCD Growth Study. Medicina (Kaunas, Lithuania). 2018;54(6).

40.	Akseer N, Mehta S, Wigle J, Chera R, Brickman ZJ, Al-Gashm S, et al. Non-communicable diseases among adolescents: current status, determinants, interventions and policies. BMC public health. 2020;20(1):1908.
